# Impact of the COVID-19 pandemic on routine surveillance for adults with chronic hepatitis B virus (HBV) infection in the UK

**DOI:** 10.1101/2021.11.10.21265651

**Authors:** Cori Campbell, Tingyan Wang, David A Smith, Oliver Freeman, Theresa Noble, Kinga A Várnai, Steve Harris, Hizni Salih, Gail Roadknight, Stephanie Little, Ben Glampson, Luca Mercuri, Dimitri Papadimitriou, Christopher R Jones, Vince Taylor, Afzal Chaudhry, Hang Phan, Florina Borca, Josune Olza, Frazer Warricker, Luis Romão, David Ramlakhan, Louise English, Paul Klenerman, Monique I. Andersson, Jane Collier, Eleni Nastouli, Salim I Khakoo, William Gelson, Graham S Cooke, Kerrie Woods, Jim Davies, Eleanor Barnes, Philippa C Matthews

## Abstract

**Background and aims:** To determine the impact of the COVID-19 pandemic on the population with chronic Hepatitis B virus (HBV) infection under hospital follow-up in the UK, we quantified the coverage and frequency of measurements of biomarkers used for routine surveillance (ALT and HBV viral load).

**Methods:** We used anonymised electronic health record data from the National Institute for Health Research (NIHR) Health Informatics Collaborative (HIC) pipeline representing five UK NHS Trusts.

**Results:** We report significant reductions in surveillance of both biomarkers during the pandemic compared to pre-COVID years, both in terms of the proportion of patients who had ≥1 measurement annually, and the mean number of measurements per patient.

**Conclusions:** Further investigation is required to determine whether these disruptions will be associated with increased rates of adverse chronic HBV outcomes.

## Background

The Coronavirus Disease 2019 (COVID-19) pandemic, caused by the severe acute respiratory syndrome coronavirus 2 (SARS-CoV-2), has resulted in over 4 million deaths worldwide (1). This reported mortality burden quantifies deaths directly attributed to COVID-19 but does not currently capture indirect disease impact arising as a result of diverted attention and resources.

In order to progress towards elimination targets for Hepatitis B virus (HBV) infection set by the World Health Organization (WHO) Global Health Sector Strategy (GHSS) on Viral Hepatitis (2) there is an urgent need for the improvement of surveillance and treatment coverage. In 2015 the WHO estimated that <1% of individuals with HBV infection were receiving antiviral treatment, and the GHSS on Viral Hepatitis set a target goal for 2030 to increase treatment coverage to 80% in treatment-eligible individuals with chronic HBV (CHB).

Once diagnosed, most individuals with CHB require regular surveillance. Decisions to treat CHB individuals with antiviral agents are made based on clinical characteristics using algorithms that combine laboratory markers of liver inflammation (typically alanine transferase, ALT), HBV DNA viral load (VL), elastography scores, ultrasound, and occasionally liver biopsy (3–5). Biopsy is recognised as a gold standard for fibrosis and cirrhosis assessment, but is rarely performed due to its invasiveness and cost (6). This places reliance on non-invasive techniques for assessment and risk stratification of untreated patients (6,7). In those already receiving antiviral treatment, laboratory parameters are monitored to ensure that virologic suppression is achieved and maintained, and to provide surveillance for complications of infection and/or of treatment.

Regular monitoring of VL and liver enzymes is recommended, typically at intervals of 3-12 months, with enhanced surveillance among males at older age, those with biochemically active hepatitis, risk factors for hepatocellular carcinoma (HCC), complications of liver disease, recent diagnosis, or changing treatment plans (8,9). An annual follow-up schedule is typical for those in whom VL and liver enzymes are consistently low, without HBV or co-morbid complications, and with no indications for treatment.

The disruptive consequences of COVID-19 for antiviral treatment access, disease surveillance and other measures for achieving HBV elimination targets have been broadly described (10,11). However, HBV service impact has not yet been quantified on a large-scale using individual-level patient data that are representative of a specific population.

## Methods

We undertook longitudinal and cross-sectional analyses using routinely-collected individual-level secondary care data (including demographic and laboratory parameters) collected across five NHS Trusts in England by the National Institute for Health Research (NIHR) Health Informatics Collaborative (HIC), as previously described (12,13). HBV treatment data were available from three NHS Trusts.

We investigated how ALT and VL surveillance varied between pre-COVID (years 2016-2019) to COVID (year 2020 and part of 2021) in CHB patients. We calculated cumulative probabilities of undergoing ≥1 ALT and HBV DNA VL measurement each year using the Kaplan-Meier method, comparing probabilities across years using the log-rank test. We quantified the mean number of ALT and HBV DNA measurements per 100 patients, during pre-COVID and COVID years, with 95% CIs calculated using the normal distribution (whereby standard error (SE) is estimated by 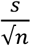 with *n* denoting sample size and *s* denoting standard deviation). We compared surveillance metrics in patients on and off antiviral therapy. Official UK government COVID incidence data are presented (14).

## Results

### (i) Coverage of ALT and VL testing (proportion tested at least once a year)

We included 3498 patients in our overall sample for 2016; this increased year on year up to 4374 patients for 2020 (**Figure S1**). In 2016, 78.6% (95% CI 77.1-79.9%) of individuals with CHB had ALT measured on ≥1 occasion throughout the year; this significantly decreased to 57.0% (95% CI 55.5-58.4%) in 2020 (**Figure 1A**). Median time to 50% of the cohort achieving ≥1 ALT measurement was <140 days for 2016-19 and 301 days for 2020. Similarly in 2016, 70.0% (95% CI 68.3-71.3%) of patients had VL measured at least once throughout the year, whilst this significantly decreased to 50.0% (95% CI 48.9-51.8%) in 2020 (**Figure 1B**). Median time to 50% of patients having achieved ≥1 VL measurement was between 158 and 167 days for 2016-19 and was 357 days for 2020. COVID incidence data are displayed in **Figure S2**.

**Figure 1.**
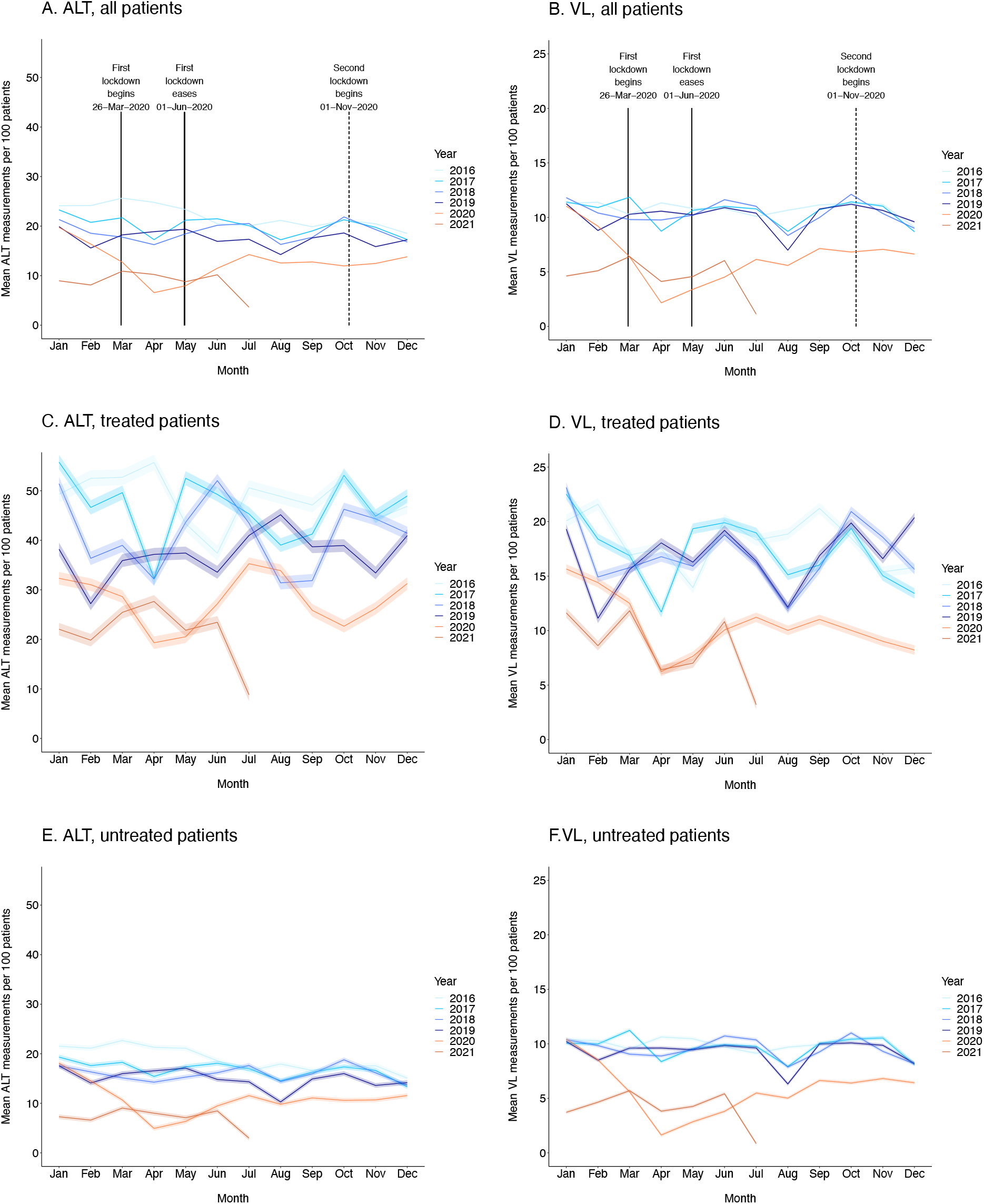
Kaplan-Meier plots demonstrating the cumulative proportion of adults with chronic HBV infection undergoing routine laboratory surveillance. Plots show patients who have had ≥1 ALT (panels A, C, E) and ≥1 HBV DNA VL (panels B, D, F) measurement each year for pre-COVID (2016-19) and COVID (2020 and part of 2021) years. For both ALT and VL, plots are stratified by treated (panels C and D, respectively) and untreated (panels E and F, respectively) patients. Dashed lines depict median time for 50% of the cohort to have the laboratory assessment undertaken. Cumulative probabilities for each year were calculated using the Kaplan-Meier method, comparing probabilities across years using the log-rank test, with 2016 serving as the reference/baseline year.

### (ii) Coverage of ALT and VL testing stratified by treatment status

A subset of patients had treatment data recorded (which remained consistent during the period observed, at 2852/3498 (81.6%) in 2016 and 3564/4374 (81.5%) in 2020). In treated patients, the proportion with ≥1 ALT measurement by the end of the year decreased from 95.2% (95% CI 85.6-98.4%) in 2016 to 74.0% (95% CI 69.7-77.6%) in 2020 (**Figure 1C**), and in untreated patients from 65.4% (95% CI 63.5-67.3%) to 53.0% (95% CI 51.1-54.8%) in 2020 (**Figure 1E**). Similarly for VL, the proportion of treated patients tested decreased from 81.0% (95% CI 68.3-88.6%) in 2016 to 69.2% (95% CI 64.8-73.1%) in 2020 among treated patients (**Figure 1D**), and from 56.7% (95% CI 54.7-58.7%) to 45.6% (95% CI 43.7-47.4%) in untreated patients (**Figure 1F**).

### (iii) Frequency of ALT and VL testing (number of tests performed each month)

Pre-COVID, the mean number of ALT measurements a month varied from 14 (95% CI 14-14) to 26 (95% CI 26-26), typically with annual dips in August and December (likely to reflect holiday periods when clinic intensity is lower) (**Figure 2A**). From March 2020 through to June 2021, this range reduced to mean number of between 7 (95% CI 7-7) and 14 (95% CI 14-14) tests per 100 patients per month, with a loss of the seasonal patterns previously observed, and reaching a nadir of 7 per 100 patients (95% CI 6-7) in April 2020 during the first lockdown. The same trend was observed for VL (**Figure 2B**), with mean numbers of VL measurements ranging from 7 (95% CI 7-7) to 12 (95% CI 12-12) per 100 patients during pre-COVID years and from 2 (95% CI 2-2) to 7 (95% CI 7-7) during the COVID period, with a nadir of 2 measurements per 100 patients (95% CI 2-2) in April 2020.

**Figure 2.**
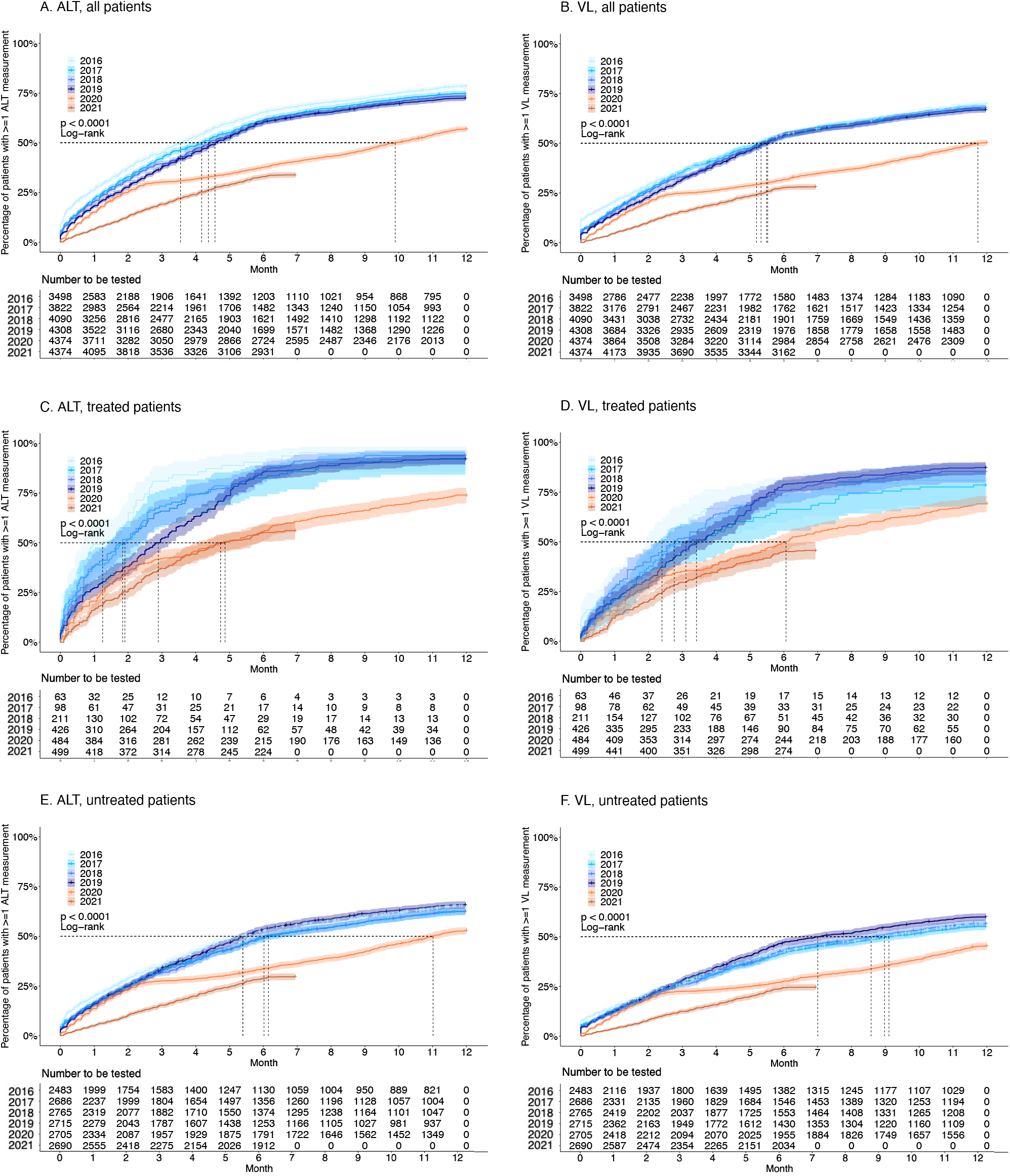
Mean numbers of ALT and HBV DNA VL measurements per 100 patients per month during pre-COVID (2016-19) and COVID (2020 and part of 2021) periods. Data are shown overall and stratified by treatment status for both VL (panels A, C, E) and ALT (panels B, D, F). Dates of UK national COVID lockdowns are denoted in overall plots. 95% CIs were calculated using the normal distribution (whereby standard error (SE) is estimated by 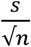 with *n* denoting sample size and *s* denoting standard deviation).

### (iv) Frequency of testing by HBV treatment status

VL and ALT measurements were more frequent in treated patients (**Fig 2 C,D**) compared to untreated patients (**Fig 2 E,F**) throughout the whole study period. In treated patients pre-COVID, the mean number of ALT measurements fluctuated between and within years but remained >25 measurements per 100 patients per month (**Figure 2C**). Mean number of monthly tests fluctuated between 19 (95% CI 18-21) and 35 (95% CI 34-37) measurements per 100 patients during the COVID period but were generally significantly lower compared to pre-COVID years. This pattern was also observed for VL, with mean numbers of measurements ranging from 11 (95% CI 11-12) to 23 (95% CI 23-24) during pre-COVID years and 6 (95% CI 6-7) to 13 (95% CI 12-13) during the COVID period (**Figure 2D**).

In untreated patients pre-COVID, mean number of ALT measurements per 100 patients per month ranged from 10 (95% CI 10-11) to 23 (95% CI 22-23), with a significant drop in the COVID period to between 5 (95% CI 5-5) and 15 (95% CI 14-15) (**Figure 2E**). This pattern was also observed for VL, with pre-COVID and COVID measurements ranging from 6 (95% CI 6-7) to 11 (95% CI 11-11) and from 2 (95% CI 2-2) to 7 (95% CI 7-7) measurements per 100 patients, respectively (**Figure 2F**).

## Discussion/Conclusions

These data from a large-scale UK clinical data source representing secondary and tertiary care centers demonstrate the negative impact of the COVID-19 pandemic on routine clinical surveillance for patients with CHB infection. Reduction in rates of surveillance closely track SARS-CoV-2 incidence (**Suppl Fig 2**) and thus periods of population lock-down.

During lockdown periods, much clinical care for viral hepatitis has been switched to telemedicine (15,16), which in most cases allows maintained contact between patients and their healthcare team, but at the detriment of laboratory monitoring. Although patients may have an option to attend a primary care or hospital phlebotomy appointment, all services have been stretched, and clinicians and/or patients may have elected to defer routine blood tests. For certain patient groups, telephone appointments are impractical, with a risk that contact may have been lost altogether during lockdown – for example in patients who do not communicate confidently in English. Furthermore, a proportion of CHB patients who left the UK early in the course of the pandemic may have been unable to return due to travel restrictions, presenting a further barrier to routine surveillance. The impact of the pandemic was seen across both treated and untreated individuals. However, pre-pandemic ALT measurements were generally more frequent in treated patients, and there was a greater impact on ALT measurement in this group during lockdowns.

We were unable to investigate how imaging-based surveillance strategies were impacted due to the COVID-19 pandemic due to the lack of complete elastography and ultrasound data in our current dataset. However, we suggest that imaging may be even more affected than laboratory surveillance as it requires the patient to attend hospital. Further collection of imaging data via the NIHR HIC will enable future investigation of this question. Disruption of both laboratory and imaging surveillance may be associated with an increased risk of CHB progression and delays in starting therapy in situations where it is indicated.

Our data represent only five centres, which are large urban secondary care providers in the South East of England. Different effects may apply in different regions or in more rural settings. Efforts and resources will be required to refine telemedicine services to optimise access to ongoing laboratory surveillance (17), to re-establish face-to-face services and to catch-up on monitoring and interventions for those who have experienced delays. Longitudinal follow-up is warranted to ensure activity is re-aligned in order to support progress to elimination goals, and investigate the relationship between these disruptions and future outcomes, including cirrhosis and hepatocellular carcinoma.

## Data Availability

This study utilised routinely-collected individual-level secondary care data (including demographic and laboratory parameters) collected across NHS Trusts in England by the National Institute for Health Research (NIHR) Health Informatics Collaborative (HIC), as previously described (1,2). The NIHR Health Informatics Collaborative (HIC) is a partnership of 29 NHS trusts and health boards, including the 20 hosting NIHR Biomedical Research Centres (BRCs), working together to facilitate the equitable re-use of NHS data for translational research. The NIHR HIC has established cross-site data collaborations in areas such as cardiovascular medicine, critical care, renal disease, infectious diseases, and cancer. All data analysed in the present study are part of the NIHR HIC database.

## Abbreviations

ALT: Alanine transferase
CHB: Chronic HBV
COVID-19: Coronavirus Disease 2019
GHSS: Global Health Sector Strategy
HBV: Hepatitis B virus
HCC: Hepatocellular carcinoma
HIC: Health Informatics Collaborative
NHS: National Health Service
NIHR: National Institute for Health Research
SARS-CoV-2: Severe acute respiratory syndrome coronavirus 2
VL: Viral load
WHO: World Health Organization

## Acknowledgments

The authors would like to thank all the research nurses and research admin staff at the contributing sites for their help in data collection and submission. The authors would also like to thank clinicians, projects managers, informaticians, and data managers at the other participating sites working to submit data.

## Ethics approval

The research database for the NIHR HIC viral hepatitis theme was approved by South Central - Oxford C Research Ethics Committee (REF Number: 21/SC/0060). All methods in this study were carried out in accordance with relevant guidelines and regulations.

