## Supplementary materials for "Impact of the COVID-19 pandemic on routine surveillance for adults with chronic hepatitis B virus (HBV) infection in the UK"

**Table of Contents**:

Figure S1. Number of HBV patients included in study period sample

Figure S2. UK COVID incidence, displayed as number of incident daily COVID cases

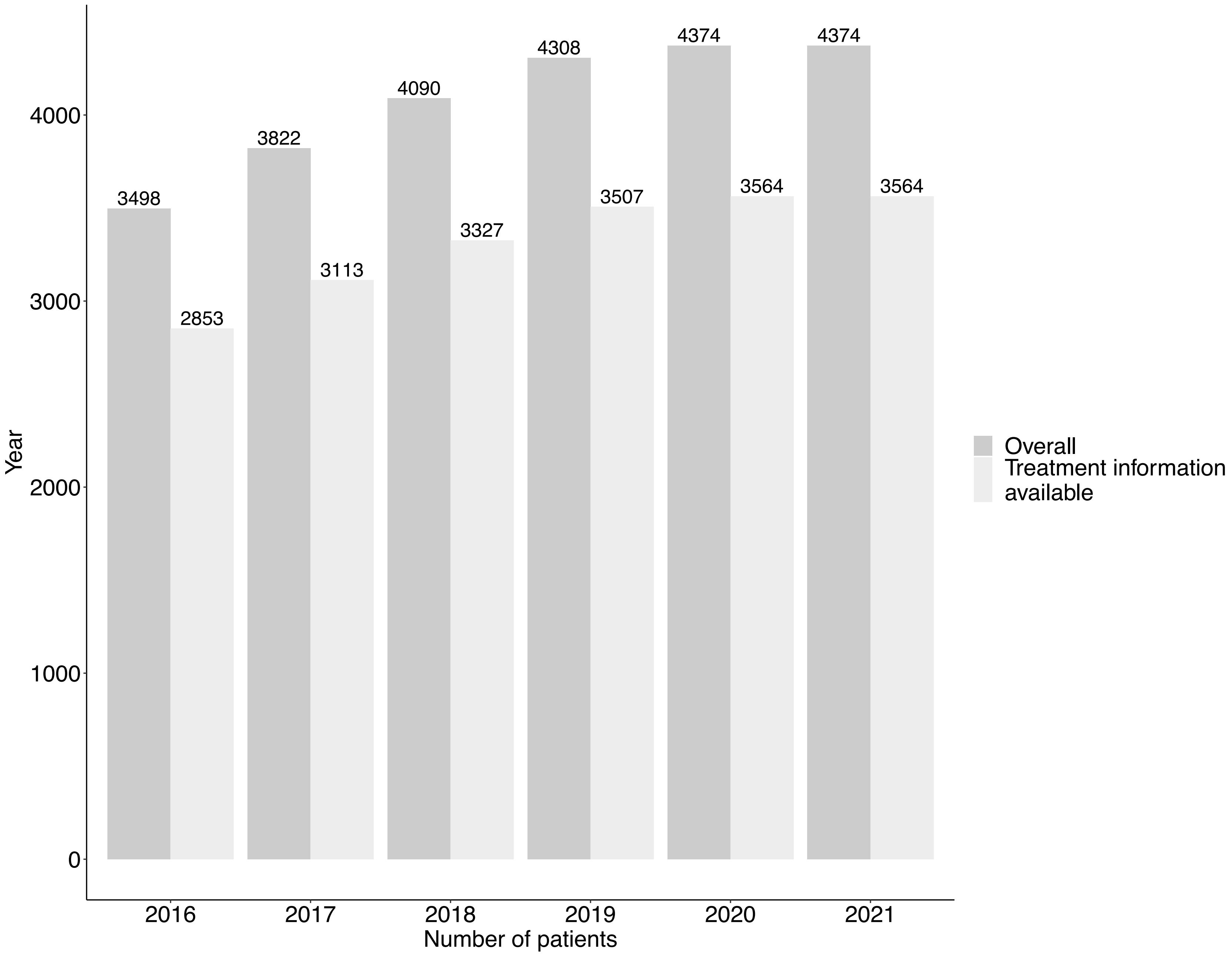

Year
0
1000
2000
3000
4000
3498
2853
3822
3113
4090
3327
4308
3507
4374
3564
4374
3564
2016
2017
2018
2019
2020
2021
Number of patients
Overall
Treatment information available

**Figure S1. Number of HBV patients included in study period sample**. Denotes number of patients overall, and those with treatment information available.

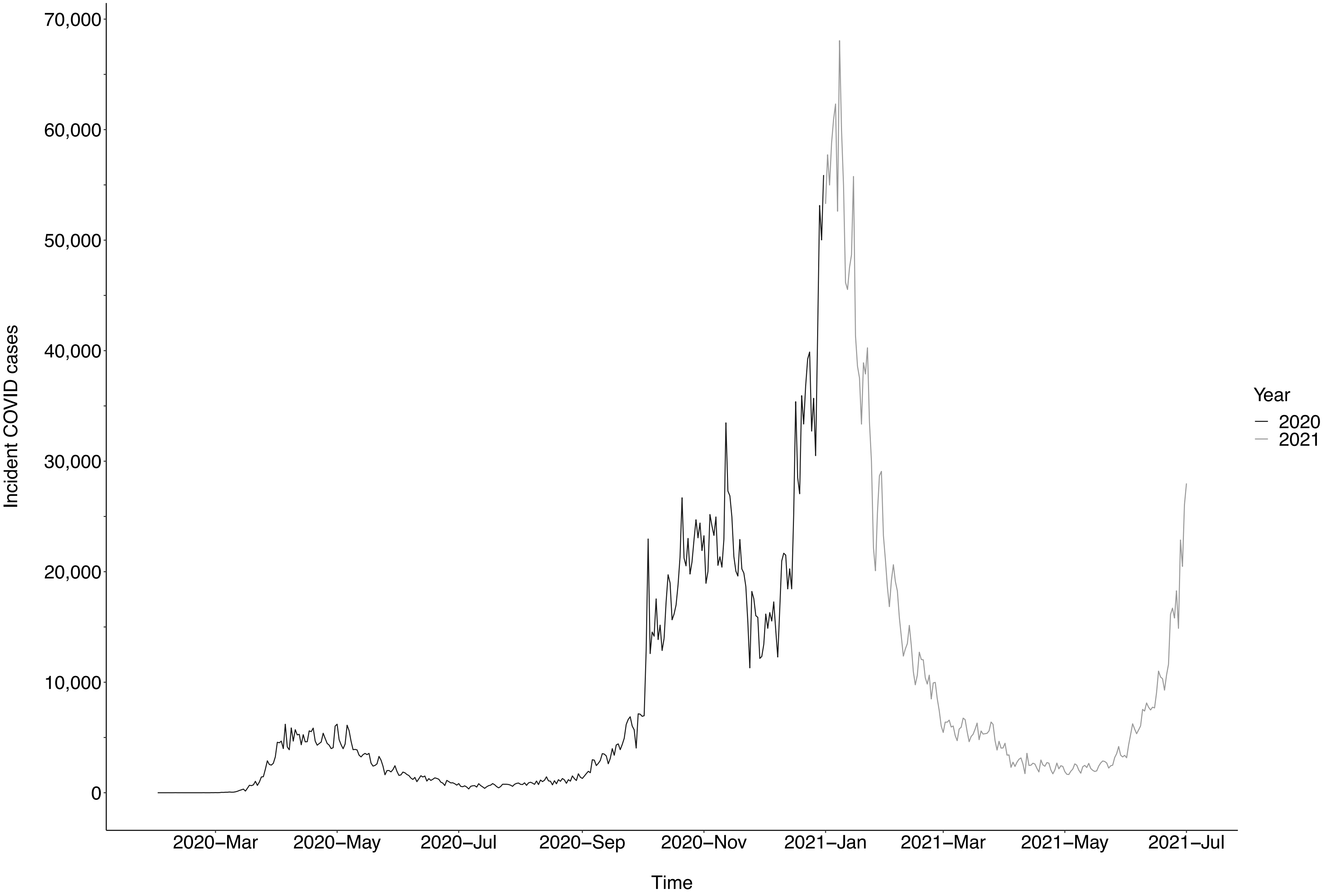


**Figure 2. UK COVID incidence, displayed as number of incident daily COVID cases**. Data from March 2020 to July 2021 are displayed.
